# Automatic detection of generalized paroxysmal fast activity in interictal EEG using time-frequency analysis

**DOI:** 10.1101/2019.12.28.19016089

**Authors:** Amir Omidvarnia, Aaron E.L. Warren, Linda J. Dalic, Mangor Pedersen, Graeme Jackson

**Author notes:** Corresponding author. Address: Medical Image Processing lab, Campus Biotech, Chemin des Mines 9, 1202 Geneva, Switzerland. (A. Omidvarnia). E-mail addresses (A. Warren), (L. Dalic), (M. Pedersen), (G.D. Jackson).

## Abstract

**Objective:** Markup of generalized interictal epileptiform discharges (IEDs) on EEG is an important step in the diagnosis and characterization of epilepsy. However, manual EEG markup is a time-consuming, subjective, and highly specialized task where the human reviewer needs to visually inspect a large amount of data to facilitate accurate clinical decisions. The objective of this study was to develop a framework for automated detection of generalized paroxysmal fast activity (GPFA), which is a characteristic type of generalized IED seen in scalp EEG recordings of patients with Lennox-Gastaut syndrome (LGS), a severe form of drug-resistant generalized epilepsy.

**Methods:** We studied 13 children with LGS who had GPFA events in their interictal EEG recordings. Time-frequency information derived from manually marked IEDs across multiple EEG channels was used to automatically detect similar events in each patient’s interictal EEG. We validated true positives and false positives of the proposed spike detection approach using both standalone scalp EEG and simultaneous EEG-functional MRI (EEG-fMRI) recordings.

**Results:** GPFA events displayed a consistent low-high frequency arrangement in the time-frequency domain. This ‘bimodal’ spectral feature was most prominent over frontal EEG channels. Our automatic detection approach using this feature identified likely epileptic events with similar time-frequency properties to the manually marked GPFAs. Brain maps of EEG-fMRI signal change during these automatically detected IEDs were comparable to the EEG-fMRI brain maps derived from manual IED markup.

**Conclusion:** GPFA events have a characteristic bimodal time-frequency feature that can be automatically detected from scalp EEG recordings in patients with LGS. Validity of this time-frequency feature is demonstrated by EEG-fMRI analysis of automatically detected events, which recapitulates the brain maps we have previously shown to underlie generalized IEDs in LGS.

**Significance:** This study provides a novel methodology that paves the way for quick, automated, and objective inspection of generalized IEDs in LGS. The proposed framework may be extendable to a wider range of epilepsy syndromes in which monitoring the burden of epileptic activity can aid clinical decision-making. For example, automated quantification of generalized discharges may permit faster assessment of treatment response and estimation of future seizure risk.

## 1 Introduction

Generalized interictal epileptiform discharges (IEDs) are a characteristic feature of the interictal (i.e., ‘between seizures’) scalp electroencephalogram (EEG) in multiple forms of epilepsy. Generalized IEDs are typically seen across bilaterally distributed scalp electrodes and show a time-varying morphology. The precise temporo-spatial appearance of these IEDs depends on the extent and location of the underlying epileptic brain network. These complex dynamics make generalized IEDs challenging to differentiate from ‘normal’ background activity, even for experienced human reviewers. Characterization of generalized IEDs in the time-frequency domain is a potential method to aid manual markup, and may assist with the development of automatic IED detection approaches.

Lennox-Gastaut syndrome (LGS) is one form of epilepsy where generalized IEDs are frequent. LGS is a severe, childhood-onset epilepsy associated with drug-resistant seizures, characteristic scalp EEG patterns, and disabling cognitive and behavioural impairments that worsen over time [Archer et al., 2014b; Intusoma et al., 2013]. Onset of LGS is typically heralded by a plateauing of cognitive development or loss of previously acquired cognitive skills, in part due to the impact of frequent epileptic activity on the developing brain [Halász et al., 2004; Warren et al., 2016].

∼8-20 Hz generalized paroxysmal fast activity (GPFA) is a characteristic of the interictal EEG in LGS. Clinical assessment of these events is currently performed manually by human experts who are required to carefully review each page of EEG over a recording that may last 24 hours or more. Although clinically informative, this process is subjective, time-consuming, and often impractical. Due to time constraints, careful manual review of the entire recording is rarely performed, with human reviewers instead ‘estimating’ the IED burden from only a sample of the total recording.

A small number of commercial ‘automated spike detection’ software programs are beginning to be used in epilepsy monitoring units (examples of such programs include ones developed by Compumedics^1^, Natus^2^, and Cadwell^3^). However, these programs were developed specifically for the detection of monomorphic *focal* IEDs as opposed to bursts of generalized discharges, which are typically more challenging for automatic detection algorithms due to their highly variable duration and morphology. Hence, these programs are not currently suitable for detection of GPFA in patients with LGS, in whom automated detection is likely to have important clinical benefits. For example, automatic quantification of GPFA burden may provide an objective and complementary measure of treatment response in patients undergoing therapeutic clinical trials [Cross et al., 2017].

In this study, we investigated EEG signal dynamics of GPFA in the time-frequency domain in a cohort of children with LGS. We showed that GPFA displays a consistent spectral power ‘bimodality’. To evaluate the utility of this feature, we searched for segments of patients’ EEG recordings with similar time-frequency properties. This was achieved by developing a novel automatic IED detection algorithm based on band-pass filtering of scalp EEG electrodes within two frequency intervals of interest, followed by integration of their instantaneous power envelopes across all scalp electrodes. Reliability of the automatically detected events was evaluated in two ways: (*i*) we assessed true positives by comparing automatically detected GPFA events to the events detected by manual human markup; and (*ii*) we assessed biological plausibility of *<u>false</u>* positive events (i.e., automatically detected events that were not identified in the manual human markup) by investigating simultaneous EEG-fMRI brain maps derived from manually marked and automatically detected events separately: the key hypothesis here was that if the automatically detected ‘GPFA-like’ events are biologically ‘meaningful’, they should yield EEG-fMRI maps that resemble ones derived from manual human markup of IEDs.

## 2 Materials and methods

### 2.1 Patients and simultaneous EEG-fMRI acquisition

Thirteen children with LGS underwent EEG-fMRI at the Royal Children’s Hospital in Melbourne, Australia. In all cases, diagnosis of LGS was defined by (*i*) multiple seizure types, including tonic seizures; (*ii*) routine interictal EEG showing GPFA and bursts of <3 Hz slow spike-and-wave (SSW); and (*iii*) cognitive and/or behavioural impairment [Arzimanoglou et al., 2009; Warren et al., 2019]. The study was approved by the Royal Children’s Hospital Human Research Ethics Committee and all patients’ parents/legal guardians gave written consent prior to participation. Patients’ EEG-fMRI results derived from manual EEG markup are reported in [Warren et al., 2019]. Their electroclinical details are summarized here in Table 1.

**Table 1:**
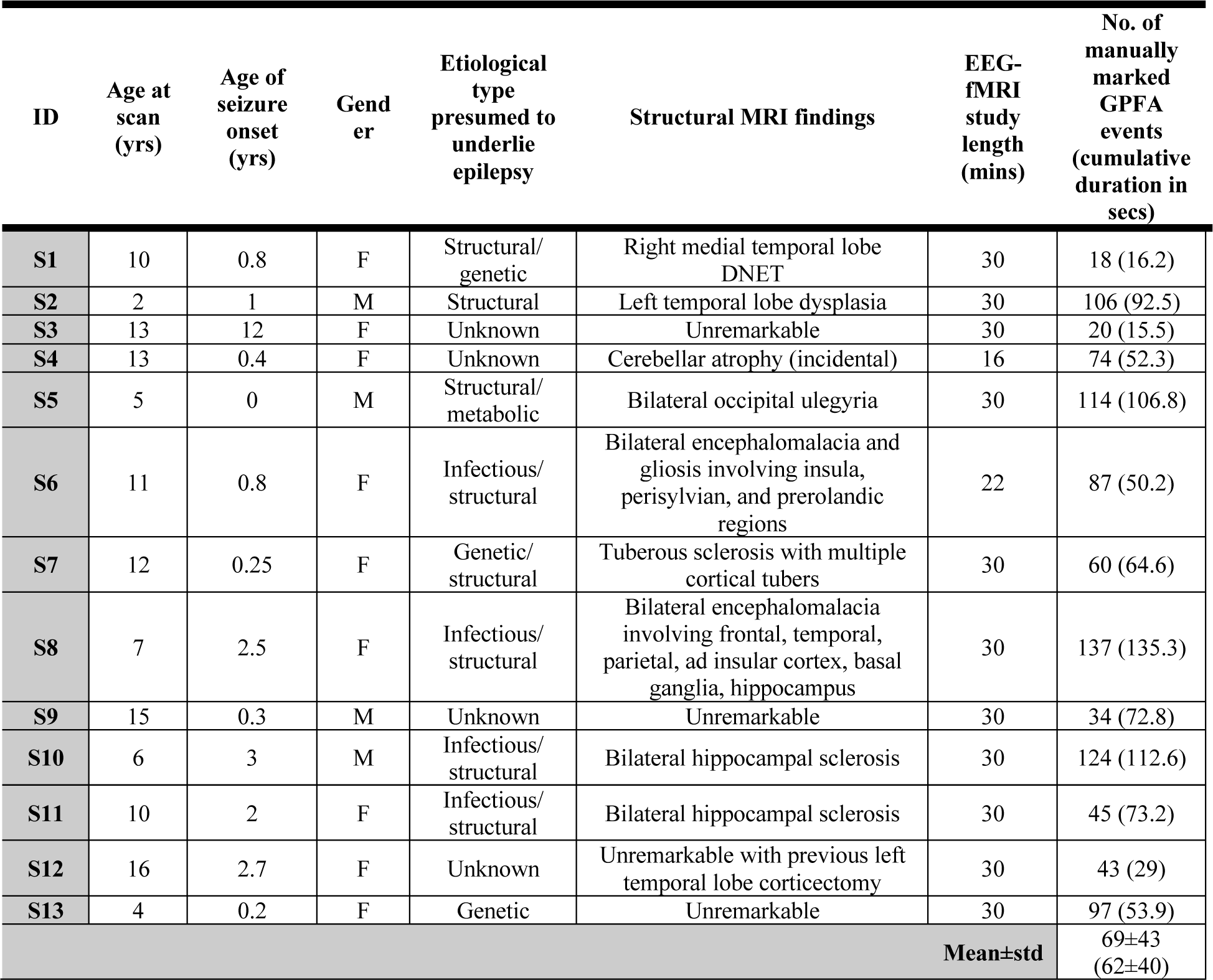
Demographic and electroclinical information of the children with LGS. For each patient, the total number and cumulative duration (in seconds) is provided for their manually marked GPFA events that were captured during EEG-fMRI scanning.

Due to intellectual disability, 12 of the 13 patients were scanned under general anaesthesia (inhalational isoflurane ≤0.8% end-tidal concentration combined with IV remifentanil ≤0.1μg/kg/min); the remaining patient (S12 in Table 1) tolerated scanning without any anaesthetic agents. fMRI data were acquired in a 3 Tesla Siemens Trio system (Munich, Germany) using an echo-planar imaging sequence with 44 interleaved slices [no gap], repetition time = 3.2 s, echo time = 40 ms and voxel size = 3.4×3.4×3.4 mm. For each patient, up to 30 minutes of fMRI was acquired (mean=28 mins). A high-resolution (0.9 mm^3^) T_1_-weighted magnetisation-prepared-rapid-gradient-echo (MPRAGE) anatomical brain image was also acquired during each session. Concurrent EEG data were acquired using a commercial 64-channel MR-compatible EEG system (Compumedics Neuroscan, Victoria, Australia) according to the 10-20 standard system. Data were recorded based on the referential montage at the sampling rate of 5000 Hz.

#### 2.1.1 Manual markup of generalized IEDs

Each patient’s in-scanner EEG was marked by an epileptologist for multiple IED types including GPFA. Table 1 summarizes the total number and cumulative duration of the GPFA event type for patients in each group. An in-scanner example of GPFA is shown in Figure 1.

**Figure 1:**
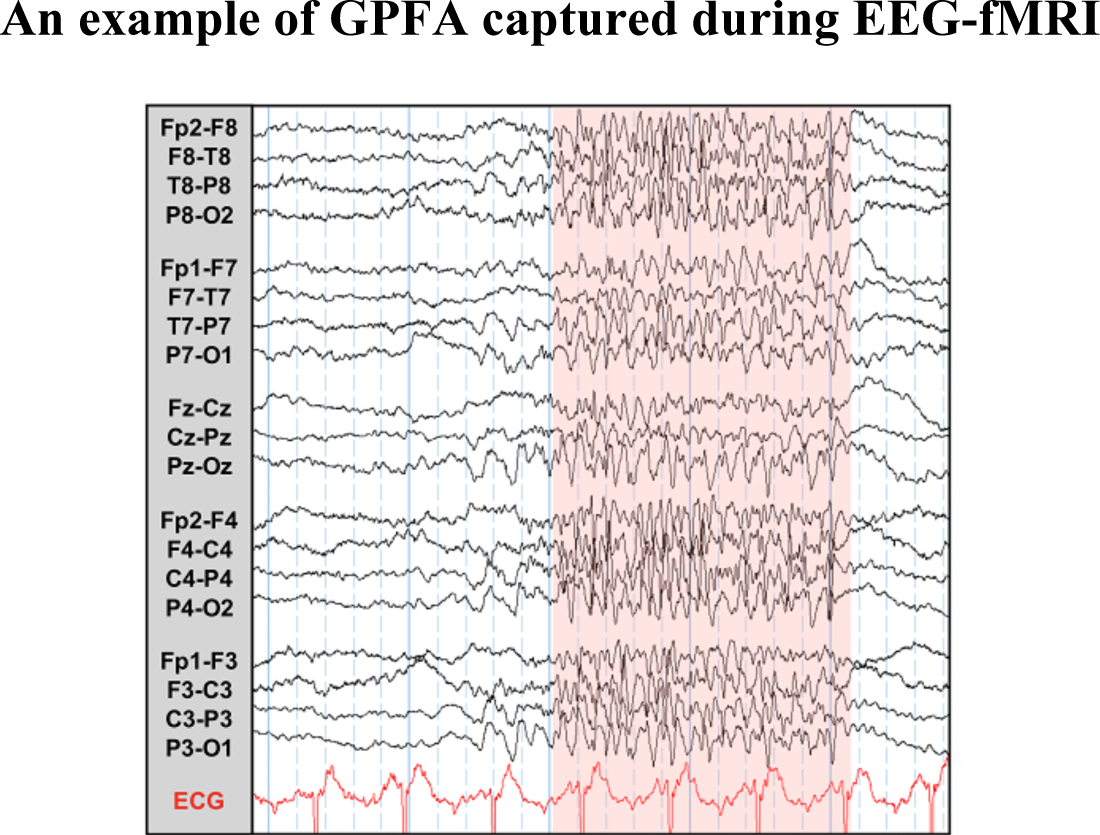
Example of a multichannel GPFA event captured during simultaneous EEG-fMRI in a patient with LGS.

### 2.2 EEG-fMRI preprocessing

fMRI data pre-processing followed our prior work [Warren et al., 2019]. Briefly, each patient’s data were corrected for slice-timing and head motion and then spatially smoothed using a Gaussian filter with full-width at half maximum of 6 mm.

EEG recordings were pre-processed using Compumedics Profusion software^4^ version 4.0 and the EEGLAB toolbox^5^ [Delorme and Makeig, 2004]. Gradient-switching and cardioballistic artifacts were corrected using average artefact template subtraction methods. To avoid computational limitations, the pre-processed EEG recordings were down-sampled to 100 Hz for further analysis.

### 2.3 Time-frequency characterization of GPFA

To quantify the time-varying properties of the manually marked GPFA events, we converted them to the time-frequency domain to reveal their joint temporal and spectral characteristics: this step was performed because the morphology of a generalized IED evolves over both time and space (i.e., scalp EEG electrodes). Additionally, its oscillations may ‘slow down’ or become ‘faster’ over the course of an individual discharge. Therefore, an appropriate quantitative detection feature for generalized IEDs must take three factors into account: (*i*) variable morphology, (*ii*) time-varying spectral content before and after the event onset, and (*iii*) multi-electrode spatial distribution. Figure 2-A and Figure 2-B demonstrate the average spectrograms of all manually marked GPFAs in a subset of scalp EEG electrodes selected from all LGS patients, calculated over a 6 s time window (−3 s to 3 s peri-onset). The selected EEG electrode in each dataset was chosen as the electrode with maximum spectral power across the frequency range of 8-20 Hz post-event onset. The relevance of this frequency band for generalized IEDs is discussed in section 3.1. In 10 out of 13 datasets, the selected electrode was located in the frontal scalp region. Visual inspection of these time-frequency representations suggested a characteristic pattern of time-frequency changes between the pre- and post-onset intervals in GPFA events. As shown in Figure 2-C and D, these pre-post event onset changes became more apparent after demeaning the time-frequency maps along the time axis (i.e., each frequency row in the maps was converted to have a mean value of zero). We observed a ‘bimodal’ burst of increased spectral power immediately following GPFA onsets that was most apparent in frontal electrodes and was seen as a simultaneous increase in spectral power in a ‘low’ frequency range of approximately 0.3-3 Hz and a ‘high’ frequency range of approximately 8-20 Hz. Based on these observations, we hypothesized a general outline for characterization of GPFA in LGS as follows:

**Figure 2:**
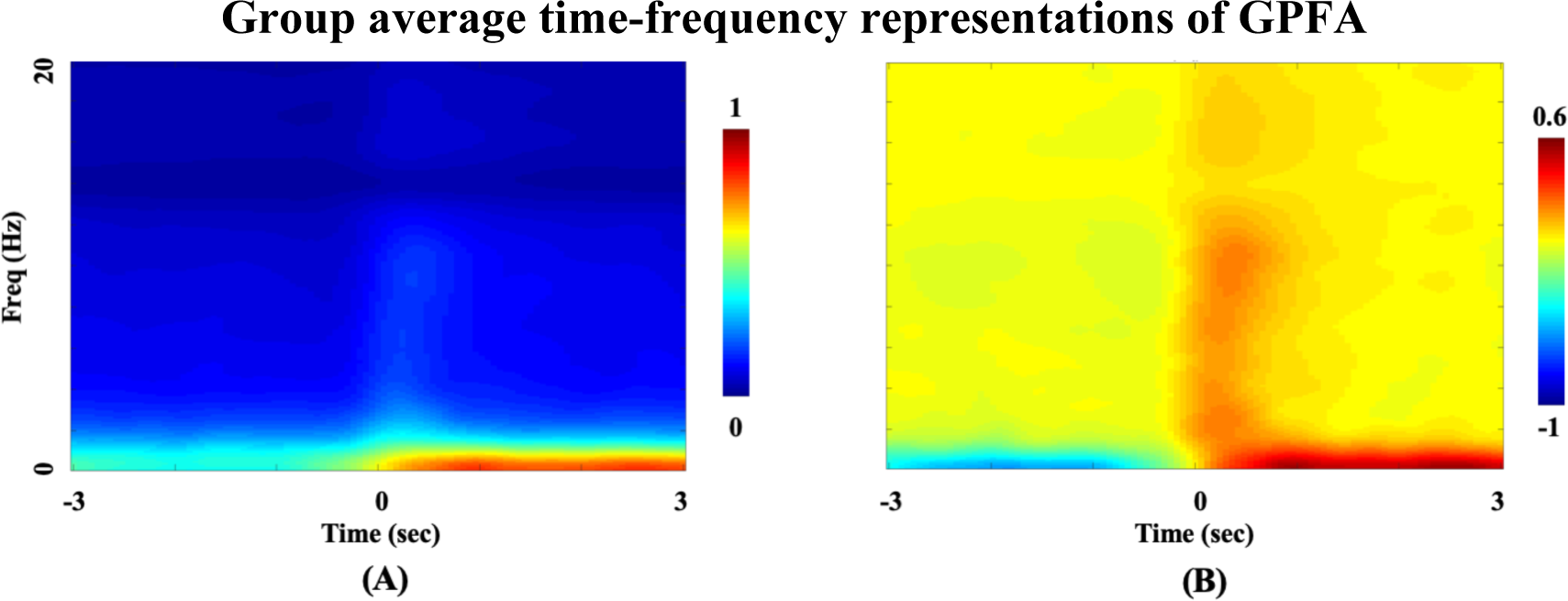
Group average time-frequency representations of GPFA events calculated in the period from 3 s before to 3 s after the onset of manually marked events over (A) all events and all patients (N=13), (B) the same events and patients as (A), but here demeaned along each frequency row (i.e., demeaning was performed along the time axis). A burst of post-onset low to high frequency activity is observed in the time-frequency maps spanning a frequency range of ∼0.3-20. This increased spectral power shows a ‘bimodal’ pattern consisting of a low-frequency patch over ∼0.3-3 Hz and a high-frequency patch over ∼8-20 Hz.

*GPFA is an interictal EEG event associated with a burst of post-onset low to high frequency activity spanning a frequency range of ∼0.3-20 Hz across multiple, particularly frontal, scalp EEG electrodes. This increased spectral power shows a bimodal pattern consisting of a low frequency patch over ∼0.3-3 Hz and a higher frequency patch over ∼8-20 Hz.*

To test this hypothesis, we considered three subject-specific parameters for each IED type in the time-frequency domain: (*i*) a post-onset low-frequency band (Δ_*LF*_ in Hz), (*ii*) a post-onset high-frequency band (Δ_*HF*_ in Hz), and (*iii*) a subset of EEG electrodes with maximal high-frequency information over Δ_*HF*_. The peri-onset interval was defined as one second before to two seconds after each manually marked IED onset time. To estimate the upper limit of Δ_*LF*_ in each patient, we divided the mean time-frequency representation of each IED type into pre- and post-onset regions. We then projected the two time-frequency planes along the frequency axis and selected the upper cut-off frequency of Δ_*LF*_ as the frequency at which the derivative of their difference changed from negative to positive. To estimate the mid-frequency of Δ_*HF*_, we detected the peak of consistent spectral ‘bumps’ above Δ_*LF*_ before and after the onset. We defined the extent of Δ_*HF*_ as the full width at half maximum of the detected peak. This analysis was performed for each patient separately.

### 2.4 Validation of the time-frequency feature via automatic IED detection

To validate the utility of the identified time-frequency feature in automatically detecting GPFA events, we searched for EEG segments with similar bimodal spectral behaviour throughout the whole of each patients’ in-scanner EEG recording. The rationale here was that if the time-frequency arrangement of GPFA is a reliable and general feature across patients, it should occur during both manually marked GPFA segments as well as ‘GPFA-*like*’ segments that were not identified in the original manual markup (as for example may occur when a ‘subtle’ event occurs that does that pass the subjective detection threshold for a human reviewer). Hence our automatic detection procedure is likely to yield both ‘true positives’ (i.e., events which coincide with the manually marked GPFA segments) as well as ‘false positives’ (i.e., events which do not coincide with the manually marked GPFA segments but which share similar bimodal spectral behaviour). In the following sections, we describe the automatic detection procedure and explain how its true/false positives were quantified.

#### 2.4.1 Automatic detection procedure

Figure 3-A and Figure 3-B show flow diagrams of the proposed automated event detection strategy at the single-channel and multi-channel levels for a typical EEG dataset. The strategy comprises two main steps: (*i*) analysis of each EEG channel separately (Figure 3-A), and (*ii*) integration of the channel-wise analyses (Figure 3-B). At the single-channel level, band-amplitude fluctuations (*BAF*s) of each channel are extracted using the Hilbert transform. The BAF envelope of a signal *X*(*t*) is defined as the absolute value of the analytic associate of its filtered version within a frequency band Δ*f*, i.e., *X*(*t*, Δ*f*) [Omidvarnia et al., 2014]. Mathematically, it is calculated by:

**Figure 3:**
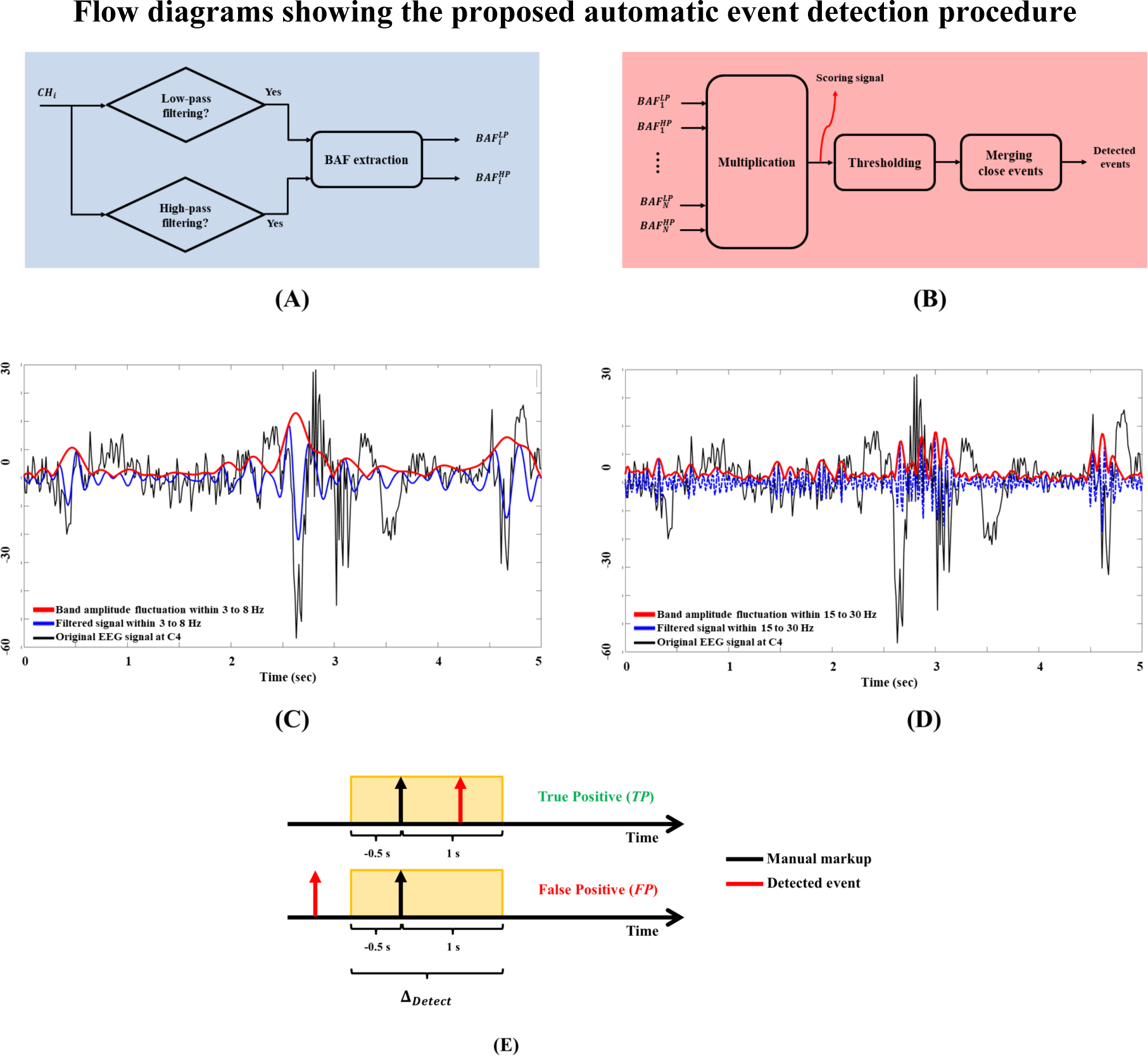
(A) Schematic of the GPFA detection at the single electrode channel level. (B) Schematic of the approach at the multi-channel level. (C) Example of a typical band amplitude fluctuation (BAF) signal for a single channel (C4) within an exemplary frequency band of 3 -8 Hz. (D) BAF envelope of the same signal extracted within a higher frequency band of 15 - 30 Hz. (E) Schematic illustrating an example of a true positive (TP) and a false positive (FP) event for the adjacency interval Δ_Detect_ of −0.5 s to 1 s peri-onset (shaded yellow rectangle). The top line illustrates a ‘true positive’ event (red arrow) with respect to Δ_Detect_ (black arrow). The bottom line shows a ‘false positive’ event (red arrow) which is outside of Δ_Detect_ (black arrow).

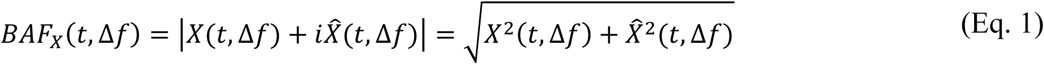

where 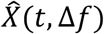 is the Hilbert transform of *X*(*t*, Δ*f*) and 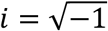. Zero-phase band-pass filtering is first performed using a Butterworth filter of order 4 in both forward and backward directions^6^. The BAF of a signal represents its time-varying spectral power within the selected frequency band Δ*f* only (i.e., BAF extraction preserves the spectral content of an event over time within a selected frequency range while excluding other frequency components that are not of interest).

Figure 3-C and Figure 3-D show, for one example dataset, the BAF envelopes for an interictal EEG segment in a typical electrode (here, C4) within a low frequency band of Δ*f* = [3 8] Hz and a high frequency band of Δ*f* = [15 30] Hz. The first step in the proposed spike detection approach is to extract two BAF envelopes for each EEG electrode within the pre-defined frequency bands Δ_*LF*_ and Δ_*HF*_. Figure 3-A illustrates the schematic of this process for a single EEG channel. For a multi-channel EEG dataset with *N*_*chan*_ number of electrodes, the per-channel BAF extraction procedure leads to 2*N*_*chan*_ BAF envelopes. The per-channel BAF signals are then integrated via multiplication over all channels at each time point: this step yields what we define here as a ‘*scoring signal’* with a duration equal to that of the entire EEG recording (red arrow in Figure 3-B). For each time-point, the scoring signal indicates the degree to which there is simultaneous high power within both Δ_*LF*_ and Δ_*HF*_ frequency bands across the intersection of all EEG channels. An increase in the scoring signal is therefore associated with the potential occurrence of a GPFA-like event with maximal information within these bands. The desired events are then detected by selecting supra-threshold time points of the scoring signal after merging adjacent events that are closer than a pre-defined time length, which we refer to as a ‘*merging window’* (here, 0.5 s). We used this merging step to avoid detecting multiple events that are closer than 0.5 s apart because such closely spaced events are more likely to represent a single GPFA burst. Section 2.4.2 explains the choice of our scoring signal threshold.

Given that obtaining Δ_*LF*_ and Δ_*HF*_ relies on an initial manual EEG markup, we first estimated these frequency bands using the manually marked datasets of our LGS cohort and then used this estimation as *a priori* knowledge for our automatic IED detection procedure. In line with the results of the group average time-frequency maps of Figure 2, we fixed Δ_*LF*_ to [0.3 3] Hz and Δ_*HF*_ of [8 20] Hz.

#### 2.4.2 True and false positive rates with reference to manual EEG markup

A key issue regarding the validity of the automatically detected events is the choice of an appropriate threshold to apply to the scoring signal, above which a potential GPFA-like event is flagged in our automatic detection procedure (Figure 3-B). To address this, we repeated the detection method for each patient’s dataset at multiple thresholds from 5% to 95% of the scoring signal amplitude in 5% increments (i.e., 19 thresholds in total per dataset) and counted the number of *true* and *false positive* events (*TP*s and *FP*s, respectively) at each threshold.

The *TP* and *FP* events were defined as follows: we considered each event (either automatically detected event or manually marked GFPA) as a Dirac delta function with zero duration and treated the set of events as a binary pulse train. We deemed an automatically detected event and a manually marked event as ‘*similar’* if their corresponding delta functions were within an *adjacency interval* Δ_*Detect*_ around the time onset of each manually marked GPFA event. Automatically detected events outside all adjacency intervals were considered to be *FP* events. Figure 3-E illustrates examples of *TP* and *FP* events with respect to a Δ_*Detect*_ of −0.5 s to 1 s peri-onset. This comparison process yielded a patient-specific *spike detection profile* for each EEG dataset where the *true positive rate* (*TPR*) is plotted across multiple scoring signal threshold levels, as in Figure 4. The threshold applied to each patient’s scoring signal was thus defined as the peak of their spike detection profile, reflecting the threshold at which the *TPR* was maximal. For each patient, the *TPR* was defined as:

**Figure 4:**
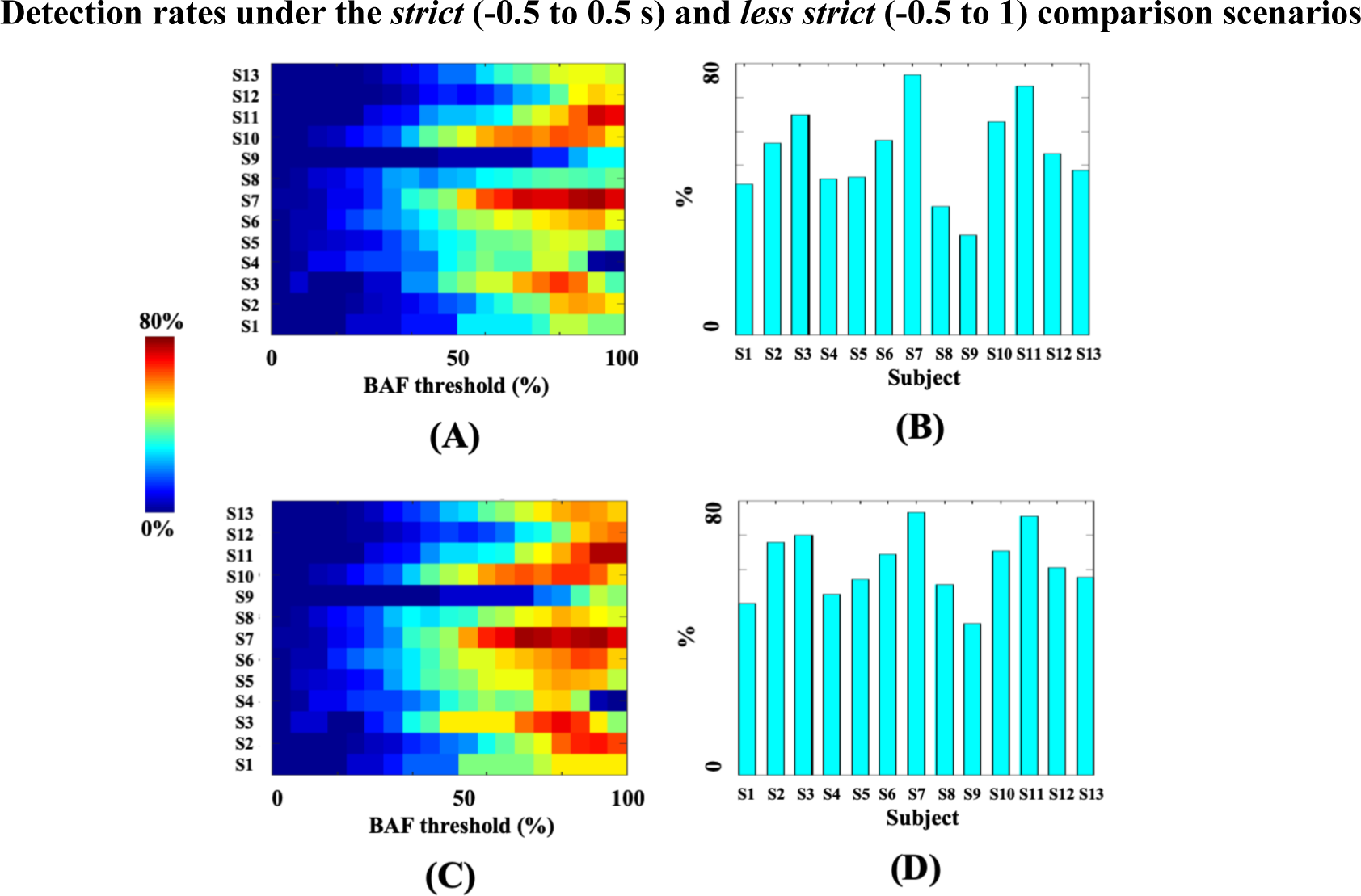
(A) and (C) Colour map of TPRs using the ‘strict’ (Δ_Detect_ = −0.5 to 0.5 s) and ‘less strict’ (Δ_Detect_ = −0.5 to 1 s) comparison procedures with multiple thresholds from 5% to 95% of the scoring signal amplitude in 5% increments. Each row in the maps represents a ‘detection profile’ for a single patient. (B) and (D): Subject-specific TPRs at the peak of the detection profile, corresponding to the panels (A) and (C), respectively. Subject IDs (S1, S2, and so on) correspond to the subject IDs in Table 1.

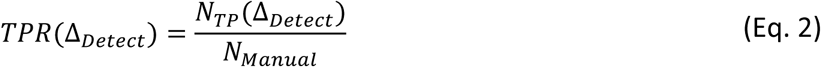

where *N*_*TP*_(Δ_*Detect*_) is the number of manually marked GPFA events which had at least one corresponding automatically detected event in the proximity of their adjacency interval and *N*_*Manual*_is the total number of manually marked GPFA events (Table 1). The *false positive rate* (*FPR*) was defined as:

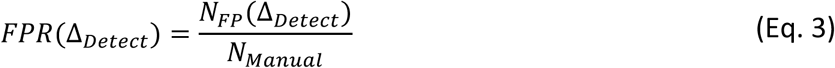

where *N*_*FP*_ is the number of automatically detected events which were outside all adjacency intervals (i.e., *N*_*FP*_(Δ_*Detect*_) = *N*_*Manual*_ − *N*_*TP*_(Δ_*Detect*_)). In this study, we used two different values for Δ_*Detect*_, i.e., −0.5 s to 0.5 s and −0.5 s to 1 s peri-onset intervals.

#### 2.4.3 Testing the biological plausibility of ‘false positives’ via EEG-fMRI analysis

Validation of *FP*s in the proposed automatic GPFA detection procedure is a non-trivial challenge. Although the *FP* events do not temporally coincide with the manually marked GPFA events (i.e., the ‘gold standard’), it is plausible that *FPs* represent genuinely epileptic events that were missed by the human expert due to, for example, markup fatigue (as is likely to occur when manually marking up lengthy EEG recordings) and/or subjective decision-making concerning the threshold at which the manually identified event is deemed to be GPFA or not. Alternatively, some proportion, or even all automatically detected events could be erroneous. In this study, we therefore chose to assess the biological plausibility of *FP*s by analysing their associated brain hemodynamic response maps using simultaneous fMRI recordings. Our rationale for this analysis was that if *FPs* are indeed biologically meaningful epileptic events, their brain hemodynamic response maps should resemble the maps derived from manually marked IEDs. A standard event-related general linear modelling (GLM) EEG-fMRI analysis approach was used [Bénar et al., 2002]. We treated the EEG events as a binary sequence of Dirac delta functions and convolved it with the canonical hemodynamic response function (HRF) available in Statistical Parametric Mapping (SPM) software, as well as its first-order derivatives with respect to time and space (i.e., a total of three event-related regressors of interest were included per GLM). As regressors of no interest, we included 6 head motion parameters in the GLM: these parameters were derived from the rigid volume realignment step performed during fMRI pre-processing. For each patient, we performed simultaneous EEG-fMRI analysis using the timing information derived from the automatically detected *TP* and *FP* events (each of these analyses were performed twice, once for each of the two Δ_*Detect*_ intervals). In addition, we repeated the analyses using the manually markup GPFA events. This led to 78 separate EEG-fMRI analyses (13 patients × 3 event types [*TP, FP*, and manually marked] × 2 adjacency intervals). For each analysis, a statistical parametric map of hemodynamic signal change was obtained by an *F-*test across the three event regressors of interest.

We hypothesized that if the detected *FP* events are non-biological, they should lead to spurious statistical parametric maps with low similarity to the maps derived from manual markup. We tested this hypothesis by calculating the pairwise spatial cross-correlation between the *F-*statistic maps associated with the *FP*s and the *F*-statistic maps associated with manual markup^7^, where a high correlation value indicates that the two maps under consideration are spatially similar. *F*-maps were compared under two circumstances: (*i*) without statistical thresholding and (*ii*) after statistical correction using an uncorrected voxel-wise *p*-value of 0.001 followed by a corrected cluster-wise *p*-value of 0.05 using Gaussian Random Field Theory [Friston et al., 1994]. All GLM analyses were performed using SPM version 12 running in MATLAB version R2016a [Veltman and Hutton, 2001].

## 3 Results

### 3.1 Estimated frequency intervals

The group-average time-frequency maps of GPFA events in Figure 2 were seen at the individual level (i.e., a bimodal spectral arrangement was observed in each patient’s interictal EEG dataset). The mean low frequency band Δ_*LF*_ was ∼ [0.3 2.7] Hz and the mean high frequency band was Δ_*HF*_ = ∼ [8.8 12.6] Hz. The high frequency band Δ_*HF*_ extended as low as [3.3 5.7] Hz in subject *S1* to [15.9 17.6] Hz in subject *S5*. Across the group, most scalp EEG channels showing the greatest high frequency power over Δ_*HF*_ were located in frontal and pre-frontal areas (10 of 13 patients). Table 2 summarizes the estimated frequency intervals (i.e., Δ_*LF*_ and Δ_*HF*_) of GPFA events for all patients.

**Table 2:** Summary of the estimated low- and high-frequency bands of GPFA for each patient.

| ID | $\Delta_{LF}$ | $\Delta_{HF}$ | Scalp EEG channel with maximum $\Delta_{HF}$ power |
| --- | --- | --- | --- |
| S1 | [0.3 1.4] Hz | [3.3 5.7] Hz | F1 |
| S2 | [0.3 2.7] Hz | [7.6 12.3] Hz | FC5 |
| S3 | [0.3 1.7] Hz | [10.8 12.6] Hz | P5 |
| S4 | [0.3 3.3] Hz | [7.9 12.2] Hz | F3 |
| S5 | [0.3 2.8] Hz | [15.9 17.6] Hz | F5 |
| S6 | [0.3 3.6] Hz | [8.4 13.6] Hz | FP1 |
| S7 | [0.3 2.6] Hz | [6.5 12.8] Hz | FP1 |
| S8 | [0.3 2.9] Hz | [9.8 13] Hz | FC1 |
| S9 | [0.3 2.7] Hz | [7.7 12.3] Hz | FP2 |
| S10 | [0.3 2.3] Hz | [8.7 12.8] Hz | PO6 |
| S11 | [0.3 2.2] Hz | [8.2 13.7] Hz | PO8 |
| S12 | [0.3 3.3] Hz | [10.2 12.8] Hz | F2 |
| S13 | [0.3 3.3] Hz | [9.4 12.4] Hz | FPZ |
| Mean | [0.3 2.7] Hz | [8.8 12.6] Hz | Frontal electrodes: 10/13 |

Note that although these frequency intervals are shown in Table 2 for each patient, the same ‘fixed’ frequency intervals (based on mean values across patients) were used for all patients the automatic detection procedure.

### 3.2 Assessing true positives via comparison with manual markup

Using the mean high/low frequency bands estimated across all patients (see section 3.1), we performed the proposed automatic event detection procedure (Figure 3) using the fixed frequency bands of Δ_*LF*_ = [0.3 3] Hz and Δ_*HF*_ = [8 20] Hz. For each detection, we defined *TP* and *FP* events under two scenarios: *strict* comparison with Δ_*Detect*_ = −0.5 s to 0.5 s peri-onset and *less strict* comparison with Δ_*Detect*_ = −0.5 s to 1 s peri-onset.

In general, the number of automatically detected events was considerably higher than the number of manually marked GPFA events. This was reflected in the number of *FP*s (*N*_*FP*_) compared to the number of *TP*s (*N*_*TP*_). *N*_*FP*_ varied from 367±179 *FP* detected events for the *strict* comparison scenario (i.e., Δ_*HDectect*_= −0.5 s to 0.5 s peri-onset) to 412±221 detected events for the *less strict* comparison scenario (i.e., Δ_*HDectect*_ = −0.5 s to 1 s peri-onset), while the range of *N*_*TP*_ was 39±21 and 46±26 detected events, respectively.

The detection results are summarized in Table 3. As expected, *TPRs* associated with the *less strict* comparison scenario were always higher than the *strict* comparison scenario (61.3± 9.8% versus 53.7± 13.6%).

**Table 3:** Summary of the automatic event detection results for **Δ**_**LF**_ = [0.3 3] Hz and **Δ**_**HF**_ = [8 20] Hz.

| $\Delta_{Detect} = -0.5 \text{ to } 0.5 \text{ s}$ | | | $\Delta_{Detect} = -0.5 \text{ to } 1 \text{ s}$ | | |
| --- | --- | --- | --- | --- | --- |
| $N_{FP}$ | $N_{TP}$ | $TPR\%$ | $N_{FP}$ | $N_{TP}$ | $TPR\%$ |
| $367 \pm 179$ | $39 \pm 21$ | $53.7 \pm 13.6\%$ | $412 \pm 221$ | $46 \pm 26$ | $61.3 \pm 9.8\%$ |

Figure 4 demonstrates the individual analysis results under the *strict* and *less strict* comparison scenarios separately. In the figure, the left column is associated with color-coded *spike detection profiles* extracted from the LGS datasets. Maximum *TPR* values associated with the peaks of the *spike detection profiles* are also plotted as bar plots on the right side.

### 3.3 Assessing False Positives: EEG-fMRI analysis

For each patient, we investigated the spatial correlations between the manual markup *F*-map and its corresponding *F*-map generated from (*i*) *all* automatically detected events (i.e., the combination of *TPs* and *FPs*), and (*ii*) *FPs* only.

Given that the definition of *FP* and *TP* events depends on the adjacency interval Δ_*Detect*_, we performed the correlation analysis for both Δ_*Detect*_ = −0.5 s to 0.5 s peri-onset and Δ_*Detect*_ = −0.5 s to 1 s peri-onset. For Δ_*Detect*_ = −0.5 s to 0.5 s peri-onset, the average correlation between the manual markup maps and the maps of all detected events was *r=*0.71±0.13. This correlation was somewhat reduced for the *F*-maps of the *FP* events to 0.63±0.09 (see Table 4).

**Table 4:**
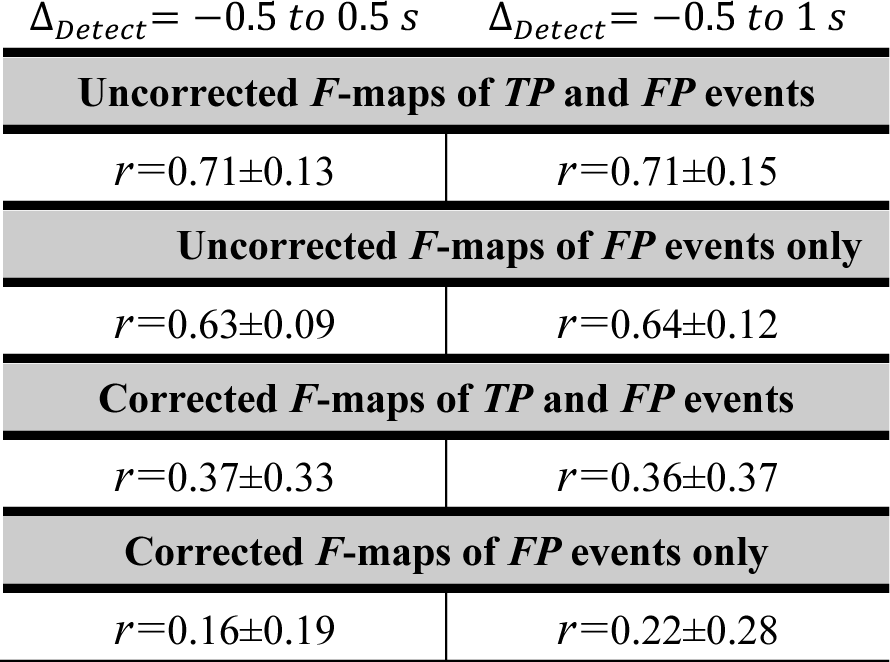
Average spatial correlation values (obtained using the FSL function fslcc) between the EEG-fMRI F-statistic maps obtained from manual markup of GPFA events versus all automatically detected events as well as false positive events only. All values are normalized between 0 (minimum spatial similarity) and 1 (full spatial overlap).

We repeated the abovementioned spatial correlation analysis for the statistically corrected *F*-maps at the voxel-wise *p*<0.001 followed by a cluster-wise *p*<0.05 correction for multiple comparisons. As Table 4 shows, the application of a statistical thresholding to the *F*-maps led to a reduction in the spatial correlation values, presumably due to fewer brain voxels that were present in the *F*-maps after the statistical correction was applied (i.e., *r=*0.37±0.33 for the maps of all detected events; and *r=*0.16±0.19 for the maps associated with the *FP* events only, all obtained at Δ_*Detect*_ = −0.5 s to 0.5 s peri-onset). As Table 4 suggests, changing Δ_*Detect*_ to −0.5 s to 1 s peri-onset did not have an obvious impact on the spatial correlation values, similar to what was observed for the uncorrected *F*-maps.

Figure 5 and Figure 6 demonstrate the subject-specific spatial correlation values as bar plots, before and after statistical correction of the *F*-maps. Examples of the GPFA *F*-maps with high correlation between automatically detected events and manual markup are illustrated in Figure 7.

**Figure 5:**
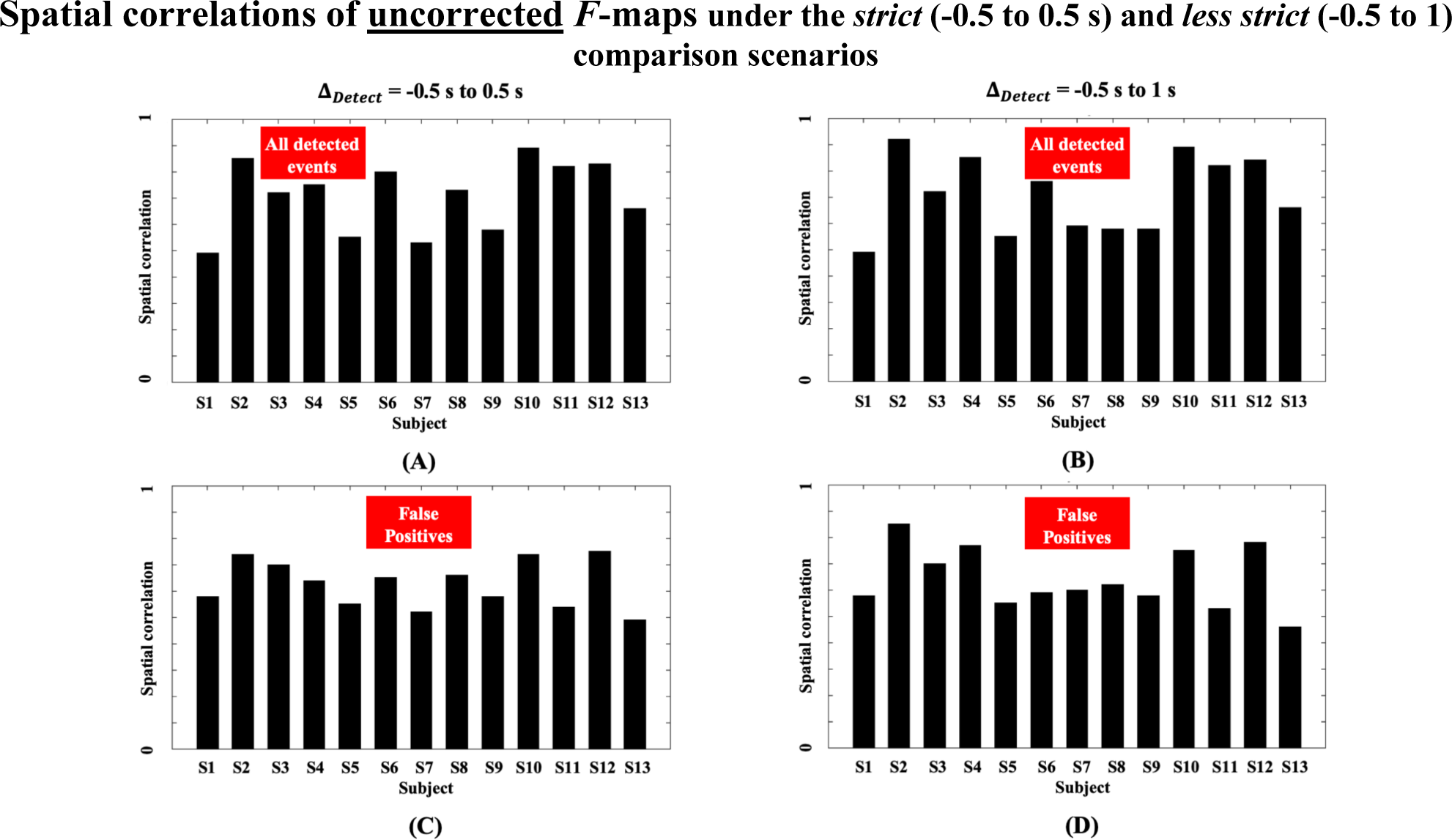
Spatial correlation values between the **<u>uncorrected</u>** F-maps of manual markup and automatically detected events for the **<u>strict comparison scenario</u>** or Δ_Detect_ = −0.5 s to 0.5 s peri-onset (panels A and C on the left) and the **less <u>strict comparison scenario</u>** or Δ_Detect_ = −0.5 s to 1 s peri-onset (panels B and D on the right). Top row panels (A and B) show correlation values with the F-maps of all detected events. Bottom row panels (C and D) show correlation with the F-maps of the false positives only. Subject IDs correspond to the subject IDs in Table 1.

**Figure 6:**
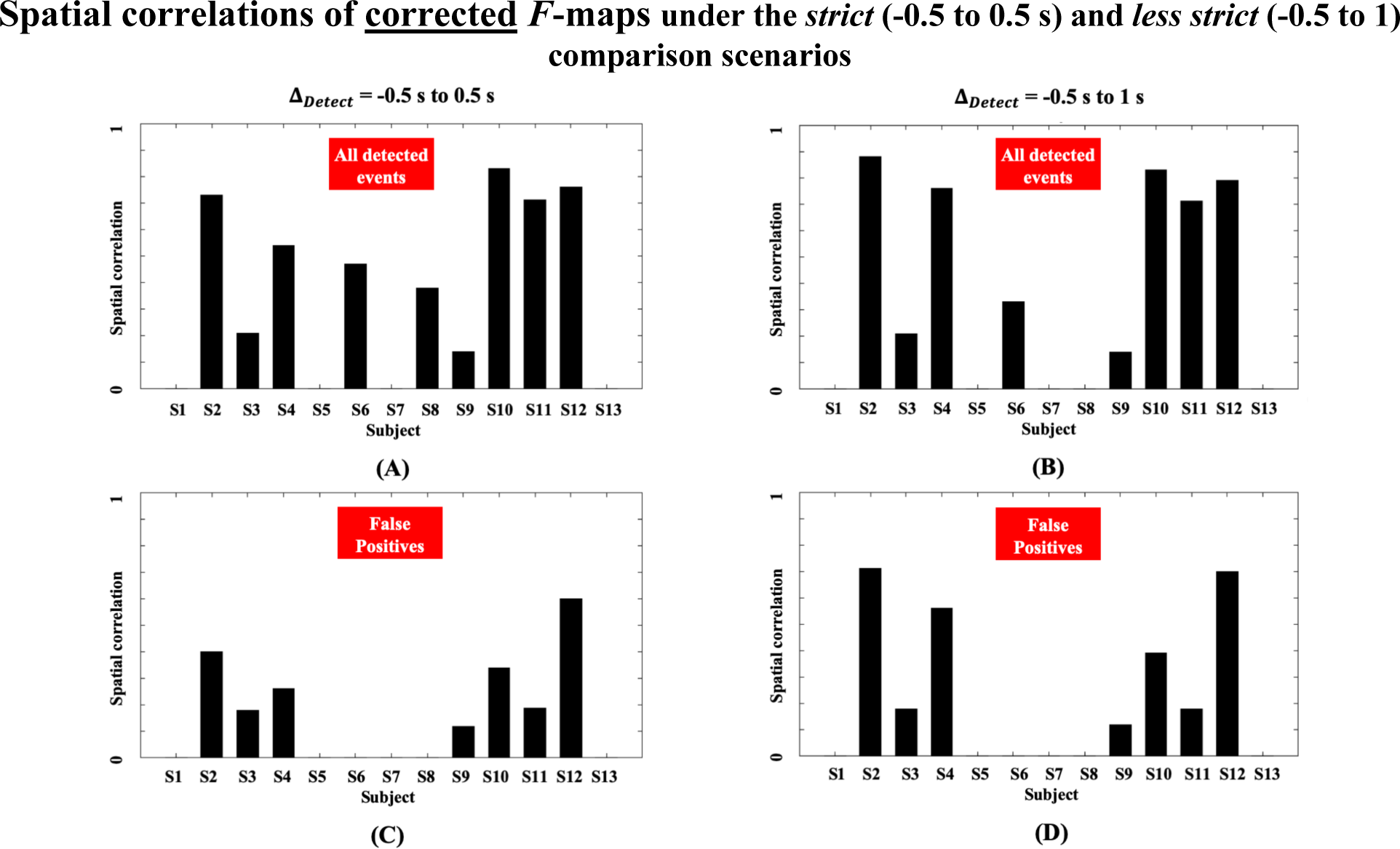
Similar caption as for Figure 5, but for the **<u>corrected</u>** F-maps.

**Figure 7:**
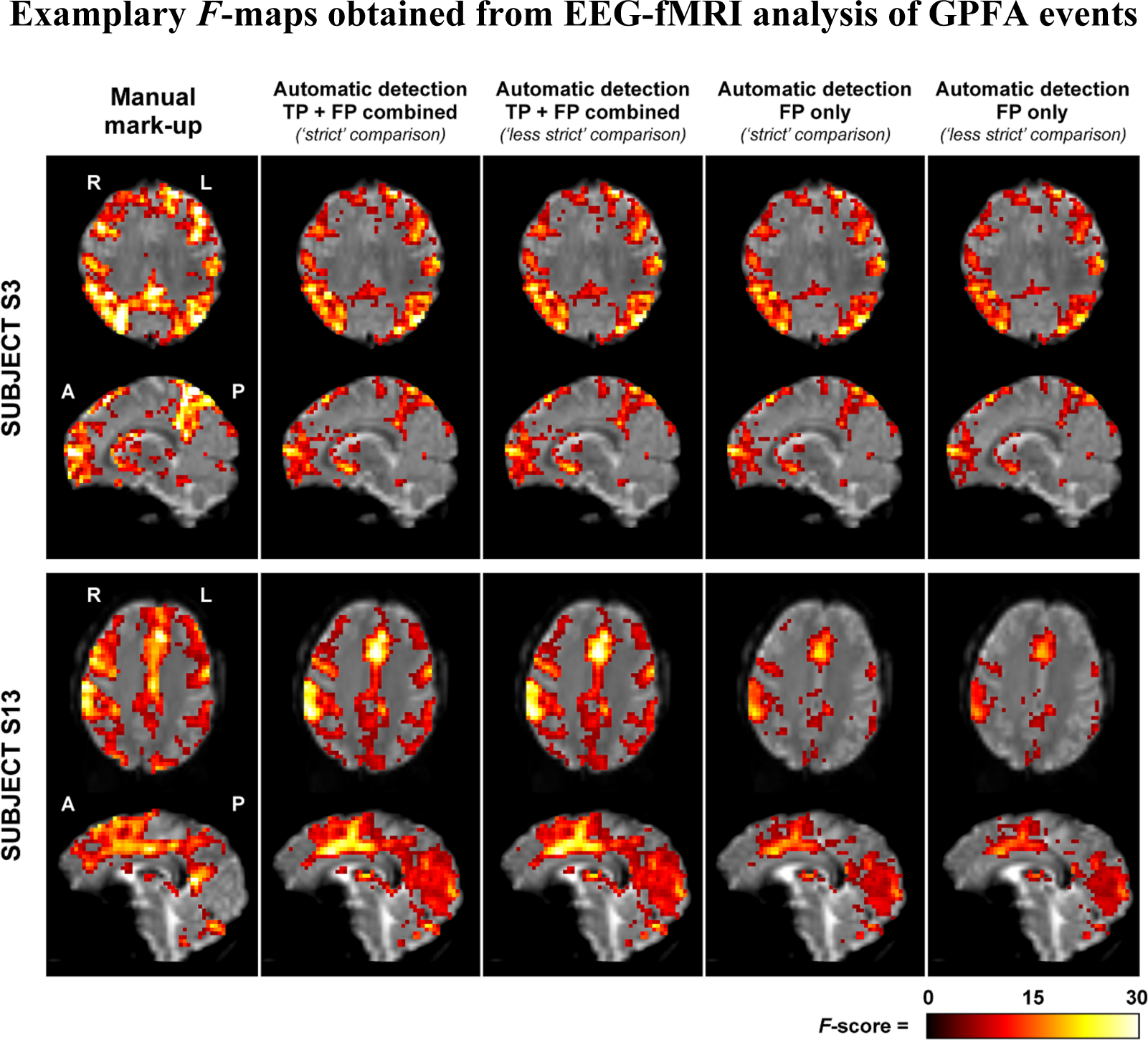
F-maps of EEG-fMRI analysis based on the timing of GPFA events for subjects S3 and S13 as two exemplary cases with high spatial agreement between automatic IED detection and manual markup. Acronyms: TP = True Positive, FP = False Positive.

## 4 Discussion

Our study reveals a bimodal time-frequency feature associated with GPFA, a generalized IED type characteristic of the interictal EEG in LGS. Our automatic IED detection approach can highlight interictal EEG segments with similar time-frequency information and can thus identify EEG events that are likely similar to manually marked IEDs. The validity of this approach is demonstrated by spatial correlation analysis showing that EEG-fMRI brain maps derived from ‘false positives’ only (i.e., automatically detected EEG events which were not originally marked up manually) are comparable to those derived from manual IED markup.

Bimodality in the spectral content of GPFA implies that these IEDs carry *nested* high frequency information that is superimposed upon slower fluctuations occurring during peri-IED intervals. This feature spans a low-frequency band of ∼ 0.3 to 3 Hz, and a high-frequency band of ∼ 8 to 12 Hz which in some patients may extend up to 20 Hz. Scalp EEG electrodes associated with frontal and pre-frontal areas tend to show the greatest high frequency power in the GPFA events, consistent with our earlier work suggesting a key role of frontal cortex in driving GPFA [Warren et al., 2019].

These results build on our previous study [Omidvarnia et al., 2018] which developed an automatic detection method for focal IEDs (i.e., events which are concentrated in time and space) in focal epilepsy patients. In contrast to this previous study, which focussed on detecting epileptic discharges with uniform morphology over time at a single EEG channel, automatic detection of generalized IEDs presents additional unique challenges: (*i*) the precise onset time of generalized discharges can be difficult to define with respect to the EEG background; (*ii*) even within a single patient, there is often variability in morphology and frequency from one discharge to the next, or over the course of a discharge train; and (*iii*) discharges can be spatially variable and show propagation over multiple EEG channels. Our current work aimed to address these challenges by inspecting generalized IEDs across multiple EEG channels and characterizing their joint temporo-spectral behaviour.

Time-frequency characterization of manually marked GPFA events suggests that their bimodal spectral behaviour is robust over patients. However, the specific frequency bands of interest may vary considerably across individuals. We assessed the robustness of this time-frequency feature via an automatic search for similar dynamics throughout patients’ interictal EEG recordings. We hypothesized that if this EEG signal feature is reliable across patients, it should be able to highlight both manually marked IEDs as well as IED-like events that were ‘missed’ in the manual markup. Across patients, the agreement between manually marked IEDs and automatically detected events varied from ∼40% to ∼80% for the two IED types. Even though the number of false positives was generally higher than the number of manually marked IEDs (see the *N*_*FTP*_ columns in Table 3), their associated statistical parametric maps generated from EEG-fMRI analysis were spatially similar to the corresponding EEG-fMRI maps derived from manual markup. This spatial overlap is above *r=*0.5 in most analyses, when the zero-thresholded *F*-statistical maps are compared. It is also similar for the *F*-maps extracted from false positives only and the maps generated based on the combination of true and false positives. Spatial overlap is reduced in both IED groups after statistical correction of the EEG-fMRI *F*-maps. This is also reflected in the group-mean correlation values of GPFA in Table 4. Less strict comparison of the automatically detected events with reference to manual markup (i.e., post-onset interval of 1 s in contrast to 0.5 s in Δ_*Detect*_) had negligible impact on the spatial correlation values of the *F*-maps. This finding suggests EEG-fMRI analysis of GPFA may show some resilience to minor variability in the precise EEG onset of IEDs (either manually marked or automatically detected), perhaps due to the comparatively slow event-related hemodynamic response measured by fMRI.

The promising detection results may facilitate faster simultaneous EEG-fMRI analysis in LGS. This is important because it may assist with pre-surgical planning for LGS, for example in guiding optimal thalamic stimulation targets in patients undergoing deep brain stimulation [Archer et al., 2014a] or identifying potentially resectable epileptogenic cortical lesions.

## 5 Conclusion

GPFA shows a characteristic bimodal time-frequency feature that can be automatically detected in patients with LGS. Utility of this time-frequency feature is demonstrated by EEG-fMRI analysis of automatically detected EEG events, which recapitulates the brain network patterns we have previously shown to underlie manually marked generalized IEDs in LGS.

## Data Availability

The EEG-fMRI datasets of this study are not available to public at this stage.

## CRediT author statement

**Amir Omidvarnia:** Conceptualization, Methodology, Software, Formal analysis, Validation, Writing-Original draft preparation. **Aaron Warren**: Conceptualization, Methodology, Data curation, Validation, Writing - Review & Editing. **Linda Dalic**: Conceptualization, Data curation, Resources, Validation, Writing - Review & Editing. **Mangor Pedersen**: Conceptualization, Methodology, Validation, Writing - Review & Editing. ***Graeme Jackson***: Conceptualization, Resources, Writing - Review & Editing, Supervision.

## Acknowledgement

A.O. acknowledges financial support through the Eurotech Postdoc Programme, co-funded by the European Commission under its framework programme Horizon 2020 (Grant Agreement number 754462). This work was supported by the National Health and Medical Research Council (NHMRC) of Australia (program grant 628952). G.J. was supported by an NHMRC practitioner fellowship (1060312). A.E.L.W was supported by a post-doctoral research fellowship from the Lennox-Gastaut syndrome Foundation (www.lgsfoundation.org). The Florey Institute of Neuroscience and Mental Health acknowledges the strong support from the Victorian Government and in particular the funding from the Operational Infrastructure Support Grant. The authors acknowledge the facilities, and the scientific and technical assistance of the National Imaging Facility at the Florey Node. We thank Dr. Magdalena Kowalczyc, Dr. John Archer and Dr. Simon Harvey for assistance with patient recruitment, EEG-fMRI data acquisition and useful discussion.

## Supplementary materials

**Figure 8:**
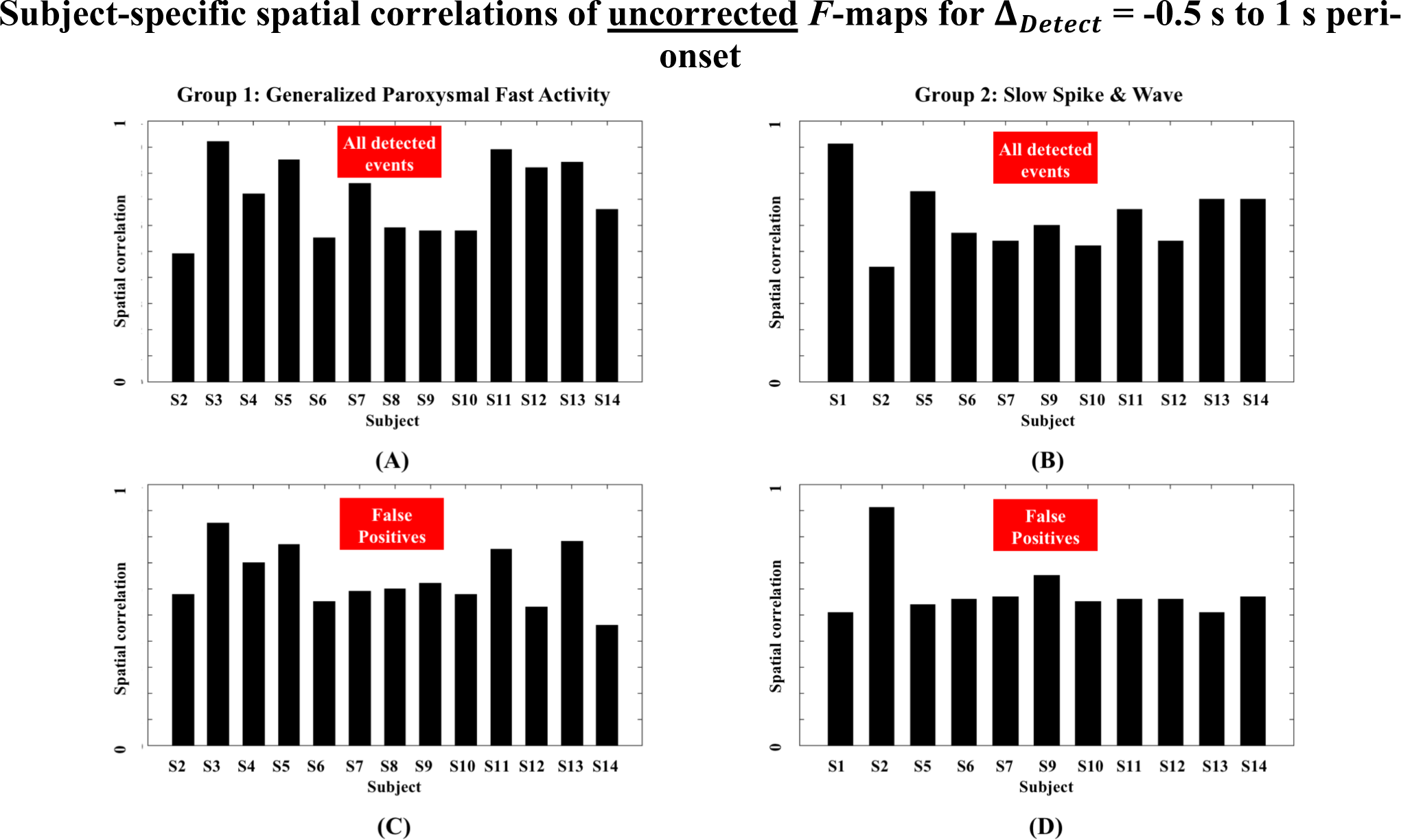
Spatial correlation values between the **<u>uncorrected</u>** F-maps of manual markup and automatically detected events for the **<u>less strict comparison scenario</u>** (Δ_Detect_ = −0.5 s to 1 s peri-onset). (A) Correlation with the F-maps of all detected events for the GPFA group. (B) Correlation with the F-maps of all detected events for the SSW group. (C) Correlation with the F-maps of the false positives only for the GPFA group. (D) Correlation with the F-maps of the false positives only for the SSW group.

**Figure 9:**
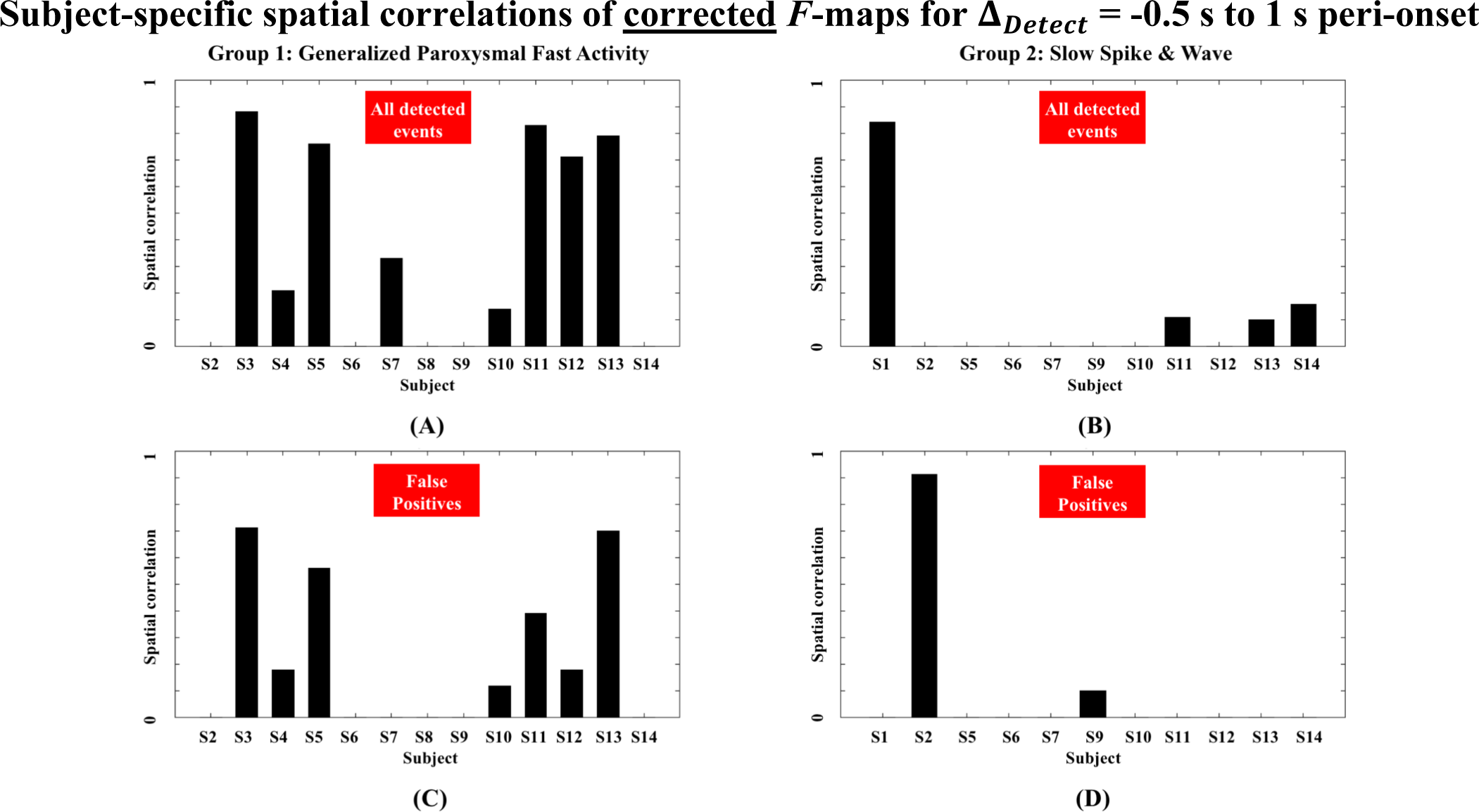
Similar caption to Figure 8, but for **<u>corrected</u>** F-maps.

## Footnotes

1 http://compumedicsneuroscan.com/curry-epilepsy-evaluation/

2 http://www.natus.com/index.cfm?page=products_1&crid=694&contentid=729

3 https://www.cadwell.com/easy3eeg/

4 https://www.compumedics.com.au/products/profusion-eeg

5 https://sccn.ucsd.edu/eeglab/index.php

6 Band-pass filtering was performed using the MATLAB command *filtfilt*. For more details, see https://www.mathworks.com/help/signal/ref/filtfilt.html

7 Spatial cross-correlation was calculated using the FSL command *fslcc*. See https://fsl.fmrib.ox.ac.uk/fsl/fslwiki/Fslutils for more details.

